# Legal Frameworks Upholding Deceased Individuals’ Rights and Enabling the Use of Cadavers in Anatomy Education and Research: A Systematic Review

**DOI:** 10.1101/2023.10.19.23297292

**Authors:** Sundip H. Charmode, Nishat Sheikh, Satish Kumar Ravi, Simmi Mehra

**Author notes:** **Corresponding author / Address:** Dr. Nishat Ahmed Sheikh Professor and Head, Department of Forensic Medicine and Toxicology, AIIMS Deoghar, Jharkhand – 814152 India, Email ID.

## Abstract

The study of human cadavers is essential for teaching, advanced training, and research in medical and anatomical sciences. Medical institutions, around the globe presently face the scarcity of cadaver supply. For the majority of countries, unclaimed bodies are still the primary source of cadavers despite guidelines issued by the International Federation of Associations of Anatomists which discourage the use of unclaimed bodies. This study aims to conduct a review of the existing national and international laws safeguarding the several rights of a deceased person. The study also reviews the existing anatomy acts (and related acts) across various countries that facilitate cadaver supply for anatomy education and research. According to PRISMA guidelines an online search was publications in four medical databases (PubMed, Scopus, Web of Sciences, and Google Scholar) was conducted from 1950 till 2022. A research review protocol was registered in PROSPERO prospectively. Using the Mesh terms like cadaver, anatomy education, dead person and rights, body donation program, unclaimed bodies, and anatomy acts. After the application of the eligibility criteria, 149 publications were shortlisted. After review of all the shortlisted articles, laws, and guidelines, using the data extraction checklist prepared by both authors, five (5) international laws; three (3) supreme court case decisions; two (2) high court case decisions; four (4) Indian penal Code Sections and 22 anatomy acts were selected. The review findings will emphasize the distinctions between India’s anatomy acts and those of other developed nations, thereby broadening our perspective as we propose a model anatomy act for uniform implementation across the country with the goal of streamlining the procurement of cadavers.

## Introduction

In the initiation of medical education curricula within medical institutions, one of the foremost and pivotal subjects that students embark upon is the discipline of human anatomy, encompassing the comprehensive study of the structural aspects of the human body ^[1]^. The foundational pillars of anatomy education and research predominantly rely on human cadaver dissection and authentic human skeletal remains. In India, the Anatomy Act, a state-level legislation akin to the Uniform Anatomical Gift Act (UAGA) in the United States, plays a pivotal role in ensuring the availability of human cadavers for anatomical education and research, while staying within the boundaries of legal provisions ^[1]^.

The previous decade has witnessed a substantial proliferation in the establishment of state-operated, central-government, and centrally autonomous medical institutions, such as AIIMS and ESIC-run medical colleges. This proliferation has led to a notable scarcity of human cadavers and genuine human skeletal specimens, which has been particularly pronounced in ESIC-run medical colleges and central government-run institutions. Presently, the primary sources of cadaver procurement primarily comprise “unclaimed” bodies, referring to individuals who have passed away without any relatives or acquaintances to claim their remains for burial, or individuals lacking the financial means to arrange for a burial. Additionally, body donation programs have emerged as a prevalent method of cadaver supply ^[2]^.

Despite the prevalence of voluntary body donation programs, which have become customary and are implemented in nearly all recently established medical institutions across the nation, there exists a substantial disparity in the number of formally announced voluntary body donations and the actual cadavers received. This disparity is influenced significantly by factors such as religious considerations, psychological barriers, and familial reasons ^[3]^. Consequently, medical institutions have turned to non-governmental organizations, such as Dadhichi Deh Dan Samiti and the Mohan Foundation, among others, as alternative sources for obtaining human cadaver donations.

A recent report issued by the Anatomy Department of Seth GS Medical College and KEM Hospital in Mumbai has documented a significant reduction in the proportion of unclaimed bodies within their mortuary’s cadaver inventory. This reduction has been observed to decline from 69.2% in the year 2005 to a notably lower figure of 18.18% in 2023. The report attributes this decline to a proactive approach taken by law enforcement authorities, emphasizing their efforts to diligently track down the relatives of deceased individuals ^[4]^. Historically, the primary source of cadavers for research purposes has been unclaimed bodies retrieved by the police. However, the widespread use of identification cards, such as the Aadhaar card, has streamlined the process of identifying bodies and locating next of kin, resulting in a reduction in the number of unclaimed bodies ^[4]^.

Presently, the situation has led to a disparity where state government medical colleges are inundated with an excess of cadavers, while newer central government and ESIC-run medical institutions face the challenge of procuring cadavers from external sources. Additionally, recent amendments in Indian legislation governing the allocation of burial grounds have imposed restrictions on the establishment of burial sites within the premises of medical institutions. Historically, these burial grounds have been used for interring dissected cadaver remains at the conclusion of the first year of academic sessions, with the subsequent excavation of original human bone sets. It is important to note that the Indian government has prohibited the export of human remains since 1985, and consequently, the buying or selling of original human bones or tissues is strictly illegal ^[5]^.

In recent years, several research studies, including those conducted by Susai S. et al. in 2023 and Bharambe V. et al. in 2019, have conducted comparative analyses of the various Anatomy Acts in India, highlighting significant disparities and inconsistencies ^[6,7]^. Additionally, a separate study by Lalwani R. et al. in 2020 engaged in a meticulous clause-by-clause comparison of these Anatomy Acts, solicited feedback from anatomists across the country and formulated a comprehensive model Anatomy Act intended for uniform adoption across all Indian states ^[1]^. This comparative analysis revealed substantial discrepancies among the different state Anatomy Acts in terms of their stated purpose, delineation of roles and responsibilities for stakeholders, regulation of body donation processes, procedures for handling unclaimed bodies, maintenance of records, dispute resolution mechanisms, penalties, and protocols for the disposal of deceased bodies ^[1]^.

Given the existence of these notable variations among the state Acts, there is an imminent and compelling need for a comprehensive revision to establish uniformity, as emphasized in an editorial featured in the Journal of the Anatomical Society of India, which serves as the official publication of the Anatomical Society of India. This editorial, published in 2002, underscores the imperative nature of this endeavor ^[1]^.

## Aim and study outcome

The aim of this study is to conduct a review of the existing laws safeguarding the various rights of a deceased person’s body both at the national and international levels. In addition, this study performs a review of the past, and current international laws in force in respective countries that promote the availability of cadavers for anatomical research and education. Our review findings will emphasize the distinctions between India’s anatomy acts and those of other developed nations, thereby broadening our perspective as we propose a model anatomy act for uniform implementation across the country with the goal of streamlining the procurement of cadavers. It should be noted that this review does not detail each anatomy act, human tissue act, national or international laws protecting dead humans around the globe. Rather, the review provides a thorough overview that accurately captures the essence of deceased human rights and laws facilitating cadaver supply throughout the world.

## Review

### Methods

To reduce the risk of bias in the study, the authors prepared a systematic review protocol and registered it with PROSPERO at the Centre for Reviews and Dissemination, University of York (Registration ID: CRD42023469534). The review protocol can be accessed from https://www.crd.york.ac.uk/prospero/.

### Search strategy

According to Preferred Reporting Items for Systematic Reviews and Meta-Analyses (PRISMA) an initial online search was conducted of articles published in 4 medical databases (PubMed, Scopus, Web of Science, and Google scholar) from 1950 till 2022 using the Mesh terms like ‘cadaver’, ‘anatomy education’, ‘dead person rights’, ‘body donation program’, ‘unclaimed bodies’, ‘anatomy acts’, and ‘human tissue act’. This led to 2790 publications.

### Eligibility criteria

The articles (original, review, case series, case reports, published and preprints) which satisfied the inclusion and exclusion criteria were eligible for review. The inclusion criteria were complete articles (free full text) published between January 1, 1950, and December 31, 2022; articles involving human species; articles authored by both foreign and Indian authors; anatomy acts, body donation acts, human tissue transplantation acts, guidelines for cadaver procurement issued by state and central government bodies at national and international level. The inclusion criteria also included Laws, Acts, Supreme court cases, High court cases, and Sections of the Indian penal code that safeguard the rights of a deceased human at national and international levels.

Exclusion criteria include articles published before 1^st^ January 1950 and after 31^st^ December 2022; articles accessible through abstracts only; publications focusing on topics other than anatomy acts, body donation acts, rights of deceased humans, and guidelines of cadaver procurement. There was a restriction for the non-English language of publications. Letters, Commentaries, Short communication, new letters, and likewise non peer reviewed publications were excluded. The Institutional Ethical Committee has been intimated and approval is sought even though it is not mandatory as this review deals with information that is freely available in public domain and is not related to patients, students, faculty, or staff directly or indirectly.

### Study selection

After applying inclusion and exclusion criteria, two authors (SC and NS) independently assessed all of the titles, abstracts, articles, acts, laws, sections, and guidelines found during the initial search, and relevant publications were shortlisted. All of the shortlisted full publications were downloaded and independently examined for relevant data using the data extraction checklist prepared by both authors. The authors of the articles, authorities issuing guidelines, and the methods of the investigations were not hidden from the reviewers. Any differences were settled through conversation or the involvement of a third reviewer (SR). The data required for our review study and the ignored data are mentioned in the checklist as shown in Table 1. We chose and incorporated the articles that contained this information.

**Table 1.** Checklist for data extraction from the selected articles.

| **Sl. no** | **Data collected** | **Data ignored** |
| --- | --- | --- |
| 1 | Any act/law/section/clause/advisory protecting the rights of dead human person. | Any act/law/section/clause protecting any right/s of alive human person/s. |
| 2 | Year of enactment of law/act (Serial no.1) and years of subsequent amendments. | General Protection guidelines of Populations against certain Consequences of War. |
| 3 | Any act/section/clause giving directives for decent burial for a dead human. |  |
| 4 | Any act/section/clause giving directives for search of a missing human. |  |
| 5 | Any act/section/clause giving directives for burial or cremation of dead human. |  |
| 6 | Any act/section/clause giving directives for cadaver procurement for anatomy education and research. |  |
| 7 | Any act/section/clause giving directives for body donation for anatomy education and research. |  |
| 8 | Any act/section/clause giving directives for cadaver procurement for surgery education and research. |  |
| 9 | Any act/section/clause giving directives about offences and penalties for violation of rights of dead humans. |  |

**Table 2.** Describes the existing international laws that protect the rights of a deceased human. These laws are compared regarding their year of enactment, years of amendments, chapter/section/clause mentioning the protection of dead human rights, and other related rights like the right to decent burial, etc.

| **Clause under consideration** | **Universal declaration of Human Rights** | **Declaration of the member states of the Organization of Islamic Cooperation** | **United Nations Commission on Human Rights** | **Geneva Convention** | **UN's Inter Agency Standing Committee's Operational Guidelines (IASC)** |
| --- | --- | --- | --- | --- | --- |
| **Year of enactment/ establishment** | 10<sup>th</sup> December 1998 at Paris. | In 1972, the Organization of Islamic Cooperation was founded. | The Commission, established in 1946. | Geneva Convention was instituted in 1864 and 1<sup>st</sup> convention meeting held on 22<sup>nd</sup> August 1864 | January 2011 published by: The Brookings – Bern Project on Internal Displacement |
| **Year when the rights of dead person were acknowledged.** | Dead person rights not declared. | In 19<sup>th</sup> Islamic conference a/k/a Cairo declaration on Human Rights held on 31<sup>st</sup> July to 5<sup>th</sup> August 1990, Cairo, Egypt. | Sixty first session was conducted on 14 March-22 April 2005 at Geneva. | Fourth (IV) meeting of Geneva Convention 1949 was held on 12<sup>th</sup> August 1949. | - |
| **Protection of rights of dead human** | No | Yes | Yes | yes | Yes |
| **Chapter/Section/Clause for dead body right.** | No | Article 3 (a) | Section IX, point 22, Clause C | Article 16, paragraph 2 |  |
| **Chapter/Section/Clause for decent burial of the dead.** | No | Article 4 |  | Article 130 |  |
| **Directive about Death certificate.** | No | No | No | Yes mentioned in Article 130. |  |
| **Directives for whether to cremate or bury.** | No | Only decent burial is mentioned.<br>No other directives. | Burial as per the wish of families and community. | Should be buried if possible as per the rites of the religion of the family.<br>If cremated then reason mentioned in death certificate. | Yes (Advice to bury if identity of dead is not confirmed) |

**Table 3.** Compares the international acts accepted by various countries/regions/continents facilitating the cadaver supply for anatomy education and research.

| Clause under consideration |  | Human Tissue Act, UK | Uniform Anatomical Gift Act, USA | India Anatomy act | Federal Decree Law, UAE | Africa act | Australia Act | Japan act | China act |
| --- | --- | --- | --- | --- | --- | --- | --- | --- | --- |
| Year of enactment/ establishment |  | 2004 | 1968 (State act) | 1949 (State act) | 2016 | 2014 | 1983 | 1949 | 1984 |
| Year of Amendment |  | - | 1987, 2006, 2007 | Several amendments by various states | - | 2017 | 1984, 1985, 1987, 2000, 2003, 2005, 2006, 2013 | - | 2007, 2010, 2014 |
| Purpose of Anatomical examination/research |  | Yes | Yes | yes | yes | yes | Yes | Yes | Yes |
| Purpose of Transplantation |  | Yes | Yes | No | Yes | Yes | Yes | Yes | Yes |
| Source of body | Unclaimed bodies | No | No | Yes | No | No | No | No | No |
|  | Donated bodies | Yes | Yes | Yes | Yes | Yes | Yes | Yes | Yes |
| Consent for photography of donated body and dissected parts. |  | Yes | No | No | No | No | No | Not clear | Not clear |
| Offence/Penalties in case of violation of act. |  | Yes | Yes | Yes | Yes | Yes | Yes | Yes | Yes |

### Data extraction process

The authors prepared a ‘data extraction form’ which was used to collect data and pilot-tested it in 10 sample articles. Two reviewers (SC and NS) worked separately to collect data from all of the selected publications/acts/laws that were included in the study. Disagreements were addressed through dialogue and the participation of a third independent reviewer (SR). The following features of the study were gathered: (i) the title of act/law/advisory; (ii) the research author/s (if applicable); (iii) the country of jurisdiction; (iv) the year of enactment of the act/law; and (v) the years of subsequent amendments. The following information was gathered about the rights of deceased humans and anatomy acts: the chapter/clause/section giving directives for rights of dead person; (ii) the chapter/section/clause giving directives for burial or cremation of dead human; (iii) the chapter/section/clause giving directives for search of a missing human; (iv) the chapter/section/clause giving directives for cadaver procurement for anatomy education and research; (vi) chapter/section/clause giving directives regarding establishing, and running body donation programs; (vii) chapter/section/clause giving directives regarding documentation procedure and consent of a donated body for anatomical examination; and (vii) reasons for cadaver shortage for anatomy education and research.

### Risk of Bias

While selecting the publications, using eligibility criteria, calibration between reviewers was conducted at the end of every 10 articles. If a high level of agreement (more than 90 %) is not achieved between the reviewers, the reviewers discuss their points of disagreement and review (and if required revise) the inclusion criteria. Two authors reviewed 5 selected publications separately using the data extraction form at a time. After a review of every 5 articles, both the authors discussed any discrepancies in data collected by them and whether was there any need to modify the data collection form. In this manner, the reviewers assessed the risk of bias. The risk of bias in systematic reviews (ROBIS) tool was used to assess the risk of bias in our review study ^[8]^.

## Results

### Literature search

The initial online search using Mesh terms recovered 3790 publications. After the application of search filters like human involvement, English language, and free full text the number was reduced to 746 publications. Duplicate articles (n=357) were deleted. Two reviewers (SC and NS) independently assessed each of the articles. After a citation search, 107 articles were added which were reviewed again by the same two authors separately. After the application of the eligibility criteria, 149 publications were shortlisted. Seventeen (17) publications were excluded due to irrelevant text which was outside the scope of the study. After a review of all the 149 shortlisted publications (including laws, acts, and guidelines) by the designated authors (SC and NS) using the data extraction checklist and subsequent discussions, the following articles were finally included in the study. There were five (5) international laws on human rights.; three (3) Indian supreme court case decisions; two (2) Indian high court case decisions; four (4) Indian penal Code Sections namely Section 297, Section 499, Section 503, and Section 377; one (1) National Advisory of India, 2021, one (1) THOTA act 1994 and twenty-two (22) anatomy acts or relevant acts.

## Discussion

### Rights of a dead person

#### Scenario of rights of a dead human in India

Sec 46 of Indian Penal Code defines death as the word “Death” denotes the death of a human being unless the contrary appears from the context. It does not define any characteristics or criteria of death. Instead, it simply states that the word “death” denotes, the death of a human being and not of an animal or some other living organism.

In the realm of jurisprudence, the Latin maxim “Actio personalis moritur cum persona” conveys that legal actions cease upon an individual’s death. This signifies the end of their legal capacity for engagement in proceedings. However, legal systems still respect a deceased person’s wishes. This includes ensuring proper funerals, honoring asset distribution preferences, protecting their reputation, and sometimes allowing ongoing legal cases involving the deceased. This legal recognition acknowledges the lasting impact of individuals even after their passing. Until relatively recently in India, there existed no explicit legal framework specifically designed to protect the rights of deceased individuals. However, Indian judicial authorities have consistently underscored the paramount importance of their responsibility to uphold the dignity and rights of deceased individuals ^[8]^. Furthermore, within the Indian legal landscape, a formal definition of the criteria that define a deceased person remains absent. Nonetheless, Indian legal jurisprudence has recognized and granted certain rights to deceased individuals that are akin to the rights enjoyed by the living ^[9]^.

#### International regulations for the rights of dead human bodies

International human rights laws are grounded in the fundamental principle of upholding human dignity. The following are examples of international agreements and legislation that explicitly address the respect owed to deceased individuals:

Article 3 (a) of the 1990 Cairo Declaration on Human Rights in Islam asserts that during the use of force and in armed conflicts, the mutilation of deceased bodies is strictly prohibited ^[10]^. In 2005, the UN Commission on Human Rights adopted a Resolution that, in Section IX, point 22, Clause C, underscores the significance of efforts to locate the bodies of those who have been killed. It emphasizes the importance of aiding in their recovery, identification, and reburial in accordance with the expressed or presumed wishes of the victims or the cultural practices of their families and communities ^[11]^.

Article 16 (II paragraph) of the Fourth Geneva Convention of 1949 mandates that, to the extent permitted by military considerations, each Party involved in a conflict must actively facilitate actions aimed at searching for the deceased and wounded, helping shipwrecked individuals and others in grave danger, and safeguarding them against pillage and ill-treatment ^[12]^.

Article 6 of the UN’s Inter-Agency Standing Committee’s Operational Guidelines on Human Rights and Natural Disasters, issued in March 2008, recommends the adoption of appropriate measures to enable the return of remains to the deceased’s next of kin. The guidelines suggest a preference for burial over cremation when identification is uncertain. They also advocate for measures that allow for the potential recovery of human remains for future identification and reburial, considering local religious and cultural practices and beliefs ^[13]^.

International humanitarian law, specifically Chapter XI, Article 130 (1) of the Fourth Geneva Convention of 1949, obliges detaining authorities to ensure the honorable burial of prisoners who pass away while in detention. Whenever possible, such burials should adhere to the religious rites of the deceased. States are further responsible for ensuring that graves are treated with respect, properly maintained, and clearly marked to ensure ongoing recognition ^[14]^.

Various countries have provisions within their criminal laws addressing acts of indignity toward deceased bodies. For instance, New Zealand’s Crimes Act of 1961 includes a section prescribing a two-year imprisonment penalty for those who treat deceased bodies, whether buried or unburied, with indignity. In the United Kingdom, Section 70 of the Sexual Offences Act 2003 pertains to offenses involving deceased bodies. South African law, under Section 14, specifically addresses individuals who engage in sexual acts with deceased bodies. It’s important to note that the United States of America lacks a federal law addressing these offenses, with different states having their own laws to penalize such acts.

#### National Regulations for rights of dead human bodies

Article 21 of the Indian Constitution, which enshrines the Right to Life, encompasses various aspects of an individual’s life, including the Right to Dignity. Through a series of judgments by both the Supreme Court and High Courts, this right has been extended to cover deceased individuals as well ^[8]^.

#### a. Supreme Court Cases

i. In the case of Parmanand Katara v. Union of India, 1989 (W. P. (Crl) No. 270 of 1988, SCC (4) 286), the Supreme Court emphasized the significance of upholding the dignity of deceased individuals ^[16]^.
ii. The Supreme Court reiterated this principle in the case of Ashray Adhikar Abhiyan v. Union of India, 2002 (W. P. (C) 143 of 2001), stressing the importance of maintaining and respecting the dignity of the deceased. Additionally, it extended the right to ensure that homeless deceased individuals receive a dignified cremation in accordance with their religious customs. This case also established a corresponding duty on the State to ensure the provision of a respectful cremation ^[17]^.
iii. In P. Rathinam v. Union of India, 1994 (SCC (3) 394), the scope of Article 21 was expanded to include the dignity of a person, emphasizing that the right to life entails a meaningful life and not mere existence. Furthermore, this right to dignity was extended to deceased individuals as well ^[18]^.

#### b. High Court Cases

i. In the case of S. Sethu Raja v. Chief Secretary, 2007 (W.P. (MD) No. 3888 of 2007), the Madras High Court directed government authorities to repatriate a deceased body from Malaysia so that a burial could take place at home in accordance with traditions and customs[19].
ii. Ramji Singh and Mujeeb Bhai Vs. State of U.P. & Ors, 2010 (PIL) No.-38985 of 2004) - The Allahabad High Court asserted that a person’s right to life encompasses the right of a deceased body to be treated with the same respect it would have deserved if alive. It is essential for the State to treat the corpse with dignity and resort to postmortem only when necessary ^[20]^.

#### c. Provisions under the Indian Penal Code, 1860

The IPC, 1860, includes various provisions that protect the rights of deceased persons. Section 297 safeguards against trespass on burial sites or places of funeral rites, and protects against insults to the religion of any person or indignity to a human corpse. Section 499 addresses dishonest misappropriation and conversion of property, while Section 503 deals with defamation and criminal intimidation. Notably, there is currently no specific legislation in India addressing sexual offenses against a deceased person. In May 2023, the Karnataka High Court recommended that the Central Government amend Section 377 IPC or introduce new provisions in the Indian Penal Code (IPC) to criminalize sexual intercourse with deceased bodies ^[21]^.

#### d. Transplantation of Human Organs and Tissues Act, 1994 (THOTA)

THOTA regulates the removal, storage, and transplantation of human organs and tissues for therapeutic purposes and prevents commercial dealings in human organs and tissues. It guarantees the deceased person the right to protect and preserve their human organs, tissues, or the deceased body itself from being harvested without their consent or the consent of close relatives ^[22]^.

#### e. National Advisory

On May 14, 2021, during the second wave of the COVID-19 pandemic in India, the National Human Rights Commission issued an advisory aimed at upholding the dignity and protecting the rights of the deceased. This advisory was prompted by a significant number of human fatalities during the second COVID-19 wave and the challenges associated with managing deceased bodies.

#### International laws regulating the supply of cadavers for anatomy education and research

##### United Kingdom (UK)

In the United Kingdom, up until the year 2005, the utilization of cadavers donated for the practice and study of human anatomy through dissection in medical institutions was governed by the Anatomy Act of 1984. This legislation specifically authorized the use of cadavers exclusively for the purpose of investigating and delineating topographical anatomy. Furthermore, the Anatomy Act of 1984 permitted the retention of dissected body parts, provided that this retention did not exceed 30% of the cadaver’s entirety. The remaining portion of the cadaver was required to undergo cremation within three years following the individual’s demise.

In parallel, a distinct legal framework known as the Human Tissue Act of 1961 regulated post-mortem examinations conducted to ascertain the cause of death, the retrieval and storage of body parts for pathological examination, and the examination of tissue for both diagnostic and research objectives.

However, a transformative shift occurred with the implementation of the UK Human Tissue Act of 2004, which took effect through Commencement Orders commencing in April 2005. This new act superseded and replaced the Human Tissue Act of 1961, the Anatomy Act of 1984, and the Human Organ Transplants Act of 1989.

The UK Human Tissue Act of 2004 exerted influence on two categories of licensable activities within educational institutions, including anatomical examination, storage of anatomical specimens, storage of human tissue and organs for scheduled purposes, photography of dissected body parts, and public display (e.g., the Hunterian Museum and associated services). Notably, this new act eliminated the prior limitation on the retention of dissected parts to 30 percent ^[23]^.

In 2012, the International Federation of Associations of Anatomists (IFAA) issued recommendations pertaining to the donation and study of human bodies, advocating for voluntary body donation and denouncing ethically contentious practices, such as the use of the bodies of executed individuals and unclaimed bodies ^[24]^.

Regarding the supply of bodies, body parts, or plastinated specimens to other institutions for educational or research purposes, it is emphasized that such transactions should not yield commercial gain. However, charging fees to recover actual costs, which encompass the expenses related to maintaining a body donation program and the preparation and transportation of specimens, is deemed acceptable. Notably, direct payment for human biological material itself is deemed ethically unacceptable ^[24]^.

In the context of anatomical body donation, informed consent should encompass explicit information regarding the potential use of images depicting a donor’s body or specific body parts for educational and/or research purposes ^[25]^.

##### United States of America (USA)

The Uniform Anatomical Gift Act (UAGA) is a model federal act originally formulated by the National Conference of Commissioners on Uniform State Laws (NCCUSL) in 1968. It was swiftly adopted by all states in the United States. However, in 1987, a revision and update of the UAGA occurred, but only 26 states chose to accept this updated version. Subsequently, numerous states have introduced non-uniform amendments to their anatomical gift acts, resulting in a lack of uniformity and harmony in the law. This divergence in the legal framework posed challenges for organ transplantation efforts ^[26]^.

Given the significant advancements in organ, eye, and tissue transplantation technologies and practices since 1987, there has been a substantial increase in the demand for donors. To address this growing need within the research community, a revised UAGA was promulgated in 2006, with active participation from key stakeholders representing donors, recipients, medical professionals, procurement organizations, regulators, and other relevant parties. Recognizing the conflict between the expressed wishes for end-of-life care outlined in advance health care directives and the initiation and/or continuation of life support systems in the context of prospective organ donation, an amendment was introduced in 2007 to Section 21 of the Revised UAGA (2006) ^[26]^.

In a survey conducted by Habicht JL et al. in 2018 spanning 70 countries, comprehensive data regarding the origins of human cadavers utilized in the context of anatomy education and research was systematically collected. The findings from this survey revealed that body donation programs have achieved well-established status as the primary or predominant source of cadavers in numerous regions. Specifically, this trend was observed prominently in most European countries, North American nations, certain regions of Asia, parts of South America, as well as in Australia and New Zealand. Consequently, these countries have managed to ensure a sufficient and reliable supply of cadavers to meet the demands of anatomy education and research endeavors ^[2]^.

##### Middle east countries

In a study conducted by N. Naidoo et al. in 2021, which focused on the Middle East region, it was observed that a substantial proportion of participants, specifically 70.7%, expressed reluctance toward cadaver donation. This reluctance was found to be influenced by several factors, including religious beliefs, psychological barriers, and familial considerations ^[3]^. Notably, despite the economic affluence of Arab Gulf countries, they do not have established body donation programs.

Interestingly, within the context of the Middle East, organ donation has garnered significant popularity and acceptance. For instance, the United Arab Emirates (UAE) enacted a Federal Decree Law pertaining to the Regulations of Human Organs and Tissue Transplantation in 2016 [28]. This legislation effectively governs various aspects of the organ and tissue donation process, the transplantation of organs and tissues prohibits human organ and tissue trafficking, safeguards the rights of both donors and recipients and prevents the exploitation of the needs of these individuals ^[29]^.

##### African countries

Certain African nations exhibit distinctive practices in contrast to North America and Europe concerning the acquisition or donation of bodies for scientific purposes, as their practices are deeply rooted in religious doctrines and are primarily regulated by these religious principles [30-33]. In specific African countries, such as Nigeria until 2015, the British Anatomy Act of 1832 remained in force. Subsequently, a consolidated and revised legislation known as the Africa Anatomy Act was enacted on December 31, 2017. This legislation was formulated to govern the donation, examination, and utilization of deceased individuals’ bodies or body parts for educational, scientific, research, therapeutic, or diagnostic objectives ^[34]^.

According to findings reported by Habicht et al. in 2018, a review of 14 African countries revealed varying practices with respect to the sources of bodies utilized for anatomical dissection. Eight of these countries exclusively employed unclaimed bodies (Ethiopia, Ivory Coast, Nigeria, Rwanda, Senegal, Tanzania, Uganda, Zambia), while four predominantly relied on unclaimed bodies (Ghana, Kenya, Malawi, Zimbabwe). Only one country, South Africa, primarily utilized donor bodies, and Libya solely relied on the importation of bodies from India. Notably, in three Oceania countries (Fiji, Samoa, and Solomon Islands), anatomical dissection was not employed in medical institutions ^[2]^.

The primary hindrance to the establishment of body donation programs in Africa is attributed to cultural and spiritual practices deeply embedded in African societies. It is worth noting that, to date, body donations in Africa primarily originate from white Africans, as the Black community is less inclined to participate in such programs ^[2]^. It remains uncertain whether there is a uniform anatomy act across the African continent. After a comprehensive evaluation, it becomes evident that amendments are necessary to address the existing deficiencies in the Africa Anatomy Act ^[34]^.

##### South-East Asian countries

###### Japan

In Japan, the dissection of human cadavers for the purposes of medical and dental education and research is legally permitted under the framework of the Postmortem Examination and Corpse Preservation Act (PECP), which was established in 1949. However, a significant limitation of the PECP is its lack of specific requirements regarding the utilization of cadavers in surgical education, research, or development. Consequently, such activities could potentially be interpreted as constituting the “destruction of corpses,” a violation of Article 190 of the Penal Code, which carries associated legal penalties.

This legal ambiguity has resulted in the limited adoption of Cadaver Skill Training in Japan, thereby restricting opportunities for the advancement of surgical techniques. To address this issue, in 2012, the Japan Surgical Society (JSS) and the Japanese Association of Anatomists (JAA) collaboratively formulated the “Guidelines for Cadaver Dissection in Education and Research of Clinical Medicine.” These guidelines were subsequently revised in 2018 ^[36]^. Through the implementation of these guidelines, an increasing number of anatomy departments within medical institutions in Japan have initiated the establishment and operation of “clinical anatomy labs,” which serve as valuable resources for enhancing surgical education among aspiring surgeons.

Furthermore, Japan enacted the Body Donation Act in 1983, also referred to as the ’Act on Body Donation for Medical and Dental Education.’ This legislation was put in place to regulate the donation of bodies, establish comprehensive guidelines governing the handling, utilization, and disposal of donated bodies, and ensure that such donations are made on a voluntary basis and with informed consent ^[36]^.

###### China

In 1984, China introduced the first “Provisional Regulation on the Use of Dead Bodies or Organs from Condemned Criminals,” which allowed death row prisoners to willingly donate their bodies or organs to hospitals or individuals in need. However, the specific requirements for obtaining verbal or written consent from these inmates were not clearly defined within the regulation ^[37]^.

To standardize organ transplantation practices in China, the State Council implemented the Regulation on Human Organ Transplantation in May 2007 ^[38]^. According to this regulation, individuals wishing to donate their organs for transplantation must provide written consent. Notably, at that time, there were no legal distinctions in China between organ donation by death row prisoners and the general population. As a result, this requirement for written authorization may also have applied to death row inmates, potentially permitting organ donation if they provided such written consent ^[39]^.

In a bid to combat organ trafficking-related crimes, the National People’s Congress Standing Committee enacted the Criminal Law Amendment in 2010 ^[40]^. To reduce reliance on organ transplants from prisoners and expand the pool of eligible donors, China initiated voluntary organ donation programs in select regions in 2010, later establishing them as a nationwide program in 2013 ^[41]^.

On September 1, 2013, the Provisions on Human Organ Procurement and Allocation (Interim) came into effect ^[41]^. These provisions require the submission of detailed records for each donor organ into the China Organ Transplant Response System (COTRS) to regulate and ensure equitable distribution of donated organs ^[39]^.

A pivotal development occurred during the National Organ Transplant Congress on October 30, 2014, when the use of organs from death row inmates was officially prohibited. It was proposed that, starting from January 1, 2015, China would exclusively utilize organs donated voluntarily by members of the general population ^[42]^.

###### Australia

Anatomy acts and related legislation exhibit variations across different states and territories, yet they share a common overarching objective of regulating the donation, management, and utilization of bodies for anatomical examination and research purposes. Several examples of such legislation in various Australian states and territories include the Human Tissue Act of 1983 in New South Wales, the Anatomy Act of 1958 in Victoria, the Anatomy Act of 1972 in Queensland, the Anatomy Act of 1939 in South Australia, the Anatomy Act of 1934 in Western Australia, the Anatomy Act of 1967 in Tasmania, the Human Tissue Act of 2006 in the Australian Capital Territory, and the Human Tissue Act in the Northern Territory ^[43]^.

Of particular significance is the Transplantation and Anatomy Act of 1983, which served to repeal several preceding laws, namely the Anatomy Act of 1884, the Sale of Human Blood Act of 1962, and the Transplantation of Human Tissue Act of 1974 ^[44]^.

### Recommendations

The authors propose several recommendations for reforming the existing anatomy legislation and achieving uniformity across all states in the country:

- We advocate for the amendment of the existing anatomy act to ensure uniform implementation throughout the nation.
- We suggest that the revised Anatomy Act should encompass comprehensive guidelines pertaining not only to the use of cadavers for anatomy education but also for surgical education and the training of surgeons.
- We emphasize that individuals wishing to donate their bodies for anatomical education should provide informed consent, explicitly specifying whether they consent to the photography and public display of their dissected body parts for educational purposes.
- In alignment with the 2012 directives from the International Federation of Associations of Anatomists (IFAA), we recommend that Indian anatomy acts should strictly limit the use of bodies for anatomy education and research to those voluntarily donated by individuals, thus adhering to ethical guidelines.

## Conclusions

In 2023, India surpassed China as the world’s most populous nation, boasting a vast number of medical colleges and emerging as one of Asia’s rapidly growing medical tourism destinations. India aspires to position itself as a prominent medical hub for the entire Asian region. Achieving this goal necessitates a substantial enhancement of its medical research infrastructure. A crucial initial step in this direction could involve the revision of the existing anatomy act and the implementation of a uniform anatomy act across all Indian states.

To ensure flexibility and accommodation of regional variations, the anatomy act can be retained as a state-specific regulation, with provisions allowing each state to adopt it with minor modifications tailored to their unique geographical, religious, and spiritual considerations.

The authors further recommend the enhancement or establishment of an organizational body such as the ’Anatomical Society of India’ (ASI) or the constitution of the ’Indian Association of Anatomists’ (IAA). These societies, like ASI or IAA (if established), should be granted authority to collaborate with the National Medical Council and assume a regulatory role in overseeing various activities, including cadaver procurement, the establishment of body donation cells, and the administration of body donation programs within medical institutions across the nation.

Additionally, the authors propose that the ’Anatomical Society of India’ should broaden its scope of engagement and collaborate with the ’Indian Surgical Society.’ Together, they should formulate comprehensive guidelines for cadaver dissection in the context of clinical medicine education and research. Furthermore, concerted efforts should be directed towards the establishment of ’Clinical Cadaver Labs’ within anatomy departments in medical institutions throughout India. This multi-pronged approach aims to bolster the research and educational capabilities of India’s medical landscape and align it with its aspiration to become a leading medical hub in Asia.

## Authors contributions

Conceptualization: SC. Data acquisition: SC, NS. Data analysis or interpretation: SC, NS, SR. Drafting of the manuscript: SC, NS. Critical revision of the manuscript: SR, NS, SM. Approval of the final version of the manuscript: all authors.

## Data Availability

All data produced in the present work are contained in the manuscript

---

**Figure 1.**
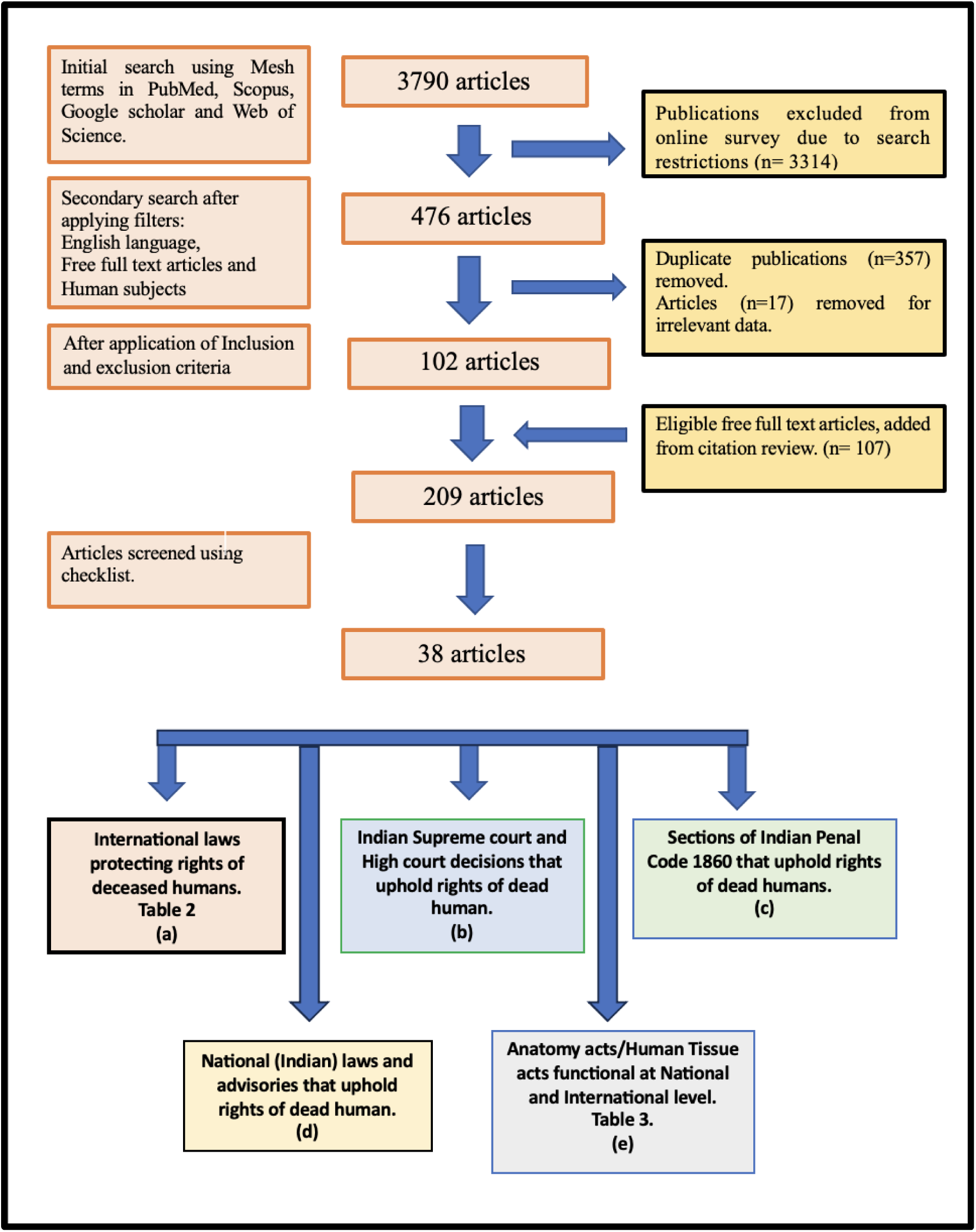
shows the Preferred Reporting Items for Systematic Reviews and Meta-Analyses (PRISMA) flowchart, which shows the step-by-step literature search and consideration/rejection procedure.

